# Determinants of acute malnutrition among children under five in arid regions, Kenya

**DOI:** 10.1101/2024.10.01.24314690

**Authors:** Dickson A Amugsi, Estelle Sidze, Faith Thuita, Valerie L. Flax, Calistus Wilunda, Linda Adair, Bonventure Mwangi, Esther Anono, Hazel Odhiambo, Stephen Ekiru, Gillian Chepkwony, Webale Albert, Monica Ng’ang’a, Joshua D. Miller, Bradley Sagara, Elizabeth Kimani-Murage, Chessa Lutter

## Abstract

Acute malnutrition, defined as a weight-for-height Z-score below -2 standard deviations of the WHO growth standards, is prevalent among children in low- and middle-income countries. Our study aimed to identify immediate, underlying, and basic determinants of acute malnutrition among children in Turkana and Samburu, two arid and semi-arid regions in Kenya. Data are from a longitudinal study that recruited children under 3 years of age, with follow-up every 4 months over six waves. Generalized estimating equations were used to assess risk factors of acute malnutrition in this population. Among immediate factors, children who recently experienced diarrhea had 19% and 23% higher odds of acute malnutrition and those who consumed animal-source foods had 12% and 22% lower odds of acute malnutrition in Turkana and Samburu, respectively. Among underlying factors, children in Turkana whose caregivers used alcohol had 32% higher odds of acute malnutrition. Among basic factors, children in Turkana whose caregivers had 3-5 or 6 or more children had 39% and 70% higher odds, whereas those in female-headed households had 34% and 81% higher odds of acute malnutrition in Turkana and Samburu, respectively. Children in Turkana’s fisherfolk communities had 36% higher odds of acute malnutrition compared with those in urban or peri-urban areas. Key risk factors for acute malnutrition included child diarrhea, caregiver’s use of alcohol (in Turkana), caregiver’s number of children, female-headed households, and fisherfolk livelihood (in Turkana), while consuming animal-source foods was associated with lower risk. Interventions should target these intersecting factors to reduce acute malnutrition in these counties.

**Key messages:**

- Acute malnutrition in children under 5 years is persistent in East Africa’s arid and semi-arid lands.
- Among immediate factors, children who had diarrhea were more likely to have acute malnutrition, while those who consumed animal-source foods were less likely to have acute malnutrition in both counties.
- Among underlying factors, children whose caregivers used alcohol were more likely to have acute malnutrition in Turkana only. Household food insecurity and water, sanitation, and hygiene were not directly associated with acute malnutrition.
- A variety of basic factors were associated with a higher likelihood of acute malnutrition, including number of children in the household, the household being headed by a female, and fisherfolk livelihood (in Turkana).

## Introduction

Acute malnutrition, commonly referred to as wasting, is a critical global health issue characterized by a child having low weight for height. This condition disproportionately impacts children under five, affecting an estimated 51.5 million children worldwide and contributing to approximately 45% of all preventable deaths in this age group (Bollinger & Trehan, 2016; Ghosh-Jerath, Singh, Jerath, Gupta, & Racine, 2017). The prevalence of acute malnutrition is highest in low- and middle-income countries, particularly in Sub-Saharan Africa and South Asia (Mena, Dedefo, & Billoro, 2018). In sub-Saharan Africa, the number of malnourished people has risen from 5.5 million to 30 million in the last decade, resulting in the deaths of more than 3.5 million children under five annually (Owolade, Abdullateef, Adesola, & Olaloye, 2022). In the Horn of Africa, over 7 million children under five are currently malnourished and in urgent need of nutrition support, with 1.9 million at risk of severe acute malnutrition-related mortality (UNICEF, 2023). Identifying risk factors of acute malnutrition has the potential to inform the development of more effective interventions to reduce its burden.

In Kenya, undernutrition – including stunting, wasting, and underweight – among children is a persistent public health challenge (KNBS & ICF, 2023; Mbogori & Muriuki, 2021). Although there has been progress toward improved child nutritional status in recent decades (Mbogori & Muriuki, 2021), the pace has been slow. For example, the prevalence of wasting decreased by only 3 percentage points in the past 30 years (Mbogori & Muriuki, 2021). At the subnational level, the situation is particularly severe in Kenya’s arid and semi-arid regions, including Turkana and Samburu counties. The prevalence of child acute malnutrition in these areas is among the highest in the country, averaging 24% (Ministry of Health, 2023) compared to the national average of 5% (KNBS & ICF, 2023). Environmental, socioeconomic, and cultural factors contribute to these observed disparities in the prevalence of acute malnutrition (Adepoju & Allen, 2019; Hawkes, Demaio, & Branca, 2017).

In Turkana and Samburu, 60% and 57% of households rely on pastoralism as their primary livelihood, respectively, which restricts access to diverse diets (Kumssa, Jones, & Herbert Williams, 2009). Limited economic opportunities, inadequate security, and lack of basic services exacerbate the vulnerability of these populations (Kumssa et al., 2009). Additionally, resource scarcity, including limited water and grazing land, often leads to violent conflicts among pastoralist communities, disrupting access to essential resources and worsening food security (McCrone, 2023). Environmental degradation due to overgrazing, deforestation, and climate change further diminishes the availability and quality of foods (Ogalo & Onyango, 2016; Okello, Ole Seno, & Nthiga, 2009). Economic constraints also limit the ability to afford nutritious diets across all livelihoods in these counties (Save the Children, 2017), despite caregivers generally being knowledgeable about infant and young child feeding recommendations (Pelto & Thuita, 2016). In addition to widespread poverty, low literacy, and historic underinvestment, Turkana and Samburu Counties also experience significant challenges with infrastructure, healthcare access, and education, each of which negatively affects child nutrition and well-being (Okello et al., 2009). Gender inequality compounds these issues, as women are often responsible for fetching water, collecting firewood, and preparing meals, leaving limited time for childcare and feeding (Christian et al., 2023). There is also limited access to safe and functional water, sanitation, and hygiene services, increasing the risk of infectious diseases, such as diarrhea, which impairs nutrient absorption (Christian et al., 2023).

Despite the complex multifaceted factors driving acute malnutrition in these counties, most studies have been cross-sectional, limiting the ability to capture dynamic changes in the risk of acute malnutrition across time. To address this gap, we conducted a 24-month longitudinal study to identify risk factors for acute malnutrition among young children in Turkana and Samburu Counties. This design allows for a more comprehensive understanding of how social, economic, and environmental factors vary and interact to influence child acute malnutrition risk. We aimed to identify key contextual factors that are associated with acute malnutrition in this region. Such information will help inform contextually relevant policies and programs to mitigate acute malnutrition in arid and semi-arid areas.

## Methods

### Study setting

This study was carried out as part of the USAID Nawiri program and was intended to contribute to program design and adaptation. The study was conducted from April 2021 to October 2023 in Turkana and Samburu counties, located in Kenya’s North Rift. These arid, resource-scarce regions are mainly inhabited by nomadic pastoralist and agro-pastoralist communities. The counties face numerous challenges, including severe droughts, limited access to essential services such as healthcare and education, and frequent intercommunal conflicts over scarce resources. Additionally, the counties have a high poverty rate and limited access to improved infrastructure (e.g., safely managed drinking water services, and improved hygiene services), hindering economic growth and human development.

### Study sample

A detailed explanation of the design, sample size estimation, and sampling strategy is described elsewhere (Wilunda et al., 2024). In brief, a multistage sampling strategy was used to ensure that findings were representative at both the county and sub-county levels. In the first stage, the population was stratified according to survey zones, as defined by respective county governments. Within each survey zone, villages were randomly selected for inclusion. Locally hired field enumerators then conducted a household listing in the selected villages to create a sample frame of households with children under three years of age. In the final stage, households were randomly chosen from this sampling frame for participation in the study. This sampling approach was designed to achieve enough completed interviews to estimate key indicators with acceptable precision. All children aged three years or younger, and their biological mothers or primary caregivers, were eligible to participate.

### Data collection

At enrollment (Wave 1), surveys were administered to collect information about household sociodemographic characteristics and individual health and nutrition behaviors and status. During subsequent survey waves, only health and nutrition information was collected.

We collected data on household characteristics, such as drinking water service, type of toilet facility, type of cooking fuel, asset ownership, and experiences with various shocks in the last four months. Household food insecurity was assessed using the eight-item Household Food Insecurity Experience Scale (Wambogo, Ghattas, Leonard, & Sahyoun, 2018). Household water insecurity was measured using the 12-item Household Water Insecurity Experiences (HWISE) Scale (S. L. Young et al., 2019).

At the individual level, eligible mothers or caregivers provided sociodemographic information, such as age, education, marital status, number of children, sex of the household head. Caregivers also reported on child feeding practices, including breastfeeding and complementary feeding on the prior day, using questions recommended by the World Health Organization and UNICEF (WHO & UNICEF, 2021). Health status was assessed by asking caregivers about common illnesses, such as diarrhea and respiratory infections, experienced by the child in the two weeks prior to the survey.

Child weight was measured using electronic Seca scales, which were designed and produced under the guidance of UNICEF. Length (for children under two years) and height were measured using measuring boards produced by Shorr Productions. Two measurements for both weight and height were taken, and the average used for analysis. Height and weight data were converted into Z-scores based on the 2006 WHO growth standards (WHO Multicentre Growth Reference Study Group, 2006).

The target sample sizes were 1,544 for Turkana and 669 for Samburu, but baseline data were collected from 1,211 participants in Turkana and 586 in Samburu. For this analysis, we included 1,201 children from Turkana and 582 from Samburu with plausible anthropometric data (weight-for-height Z-scores between -5 SD and +5 SD) and complete data for relevant covariates.

### Outcome variable

The primary outcome for this analysis was acute malnutrition. Children were categorized as being acutely malnourished if their weight-for-height Z-scores were below -2 standard deviations or not acutely malnourished if their weight-for-height Z-scores were greater than or equal to -2 standard deviations.

### Explanatory variables

Independent variables were selected based on a conceptual framework for acute malnutrition in Africa’s drylands, which categorizes contributing determinants/factors into three levels: immediate, underlying, and basic (**Supplemental Figure 1: Annex D**) (H. Young, 2020). Immediate factors included child diarrhea in the prior two weeks and consumption of breastmilk, animal-source foods (meat, fish, eggs, or dairy products), and fruits and vegetables (all types) in the prior day, each coded as dichotomous (yes/no).

Underlying factors included food insecurity, caregiver behaviors, and water, sanitation, and hygiene variables. Household food insecurity was categorized as no- to-mild and moderate-to-severe using established guidelines (Ballard, Kepple, & Cafiero, 2013). Self-reported caregiver alcohol consumption (yes/no) was considered a proxy for inadequate caregiving practices. Household water insecurity was categorized as no-to-marginal, low, moderate, or high. Other factors included water source (improved/unimproved), water storage (open container/closed container), treatment of drinking water (yes/no), self-reported caregiver handwashing after toilet use (yes/no), household open defecation (yes/no), and disposal of child stool (safe/unsafe).

Basic factors included household size (<4, 4-6, 7+ members) and household ownership of poultry (yes/no) or a television or radio (yes/no). Wealth was determined based on a factor analysis of household assets, housing materials, and access to utilities, and divided into tertiles (low, medium, high). The sex of the household head (male or female) and caregiver’s marital status (unmarried, married and only wife, married and a co-wife) were included, along with the caregiver’s parity (0-2, 3-5, 6+ children), age (<25, 25-34, 35+ years), and educational level (formal or no formal education). Respondents also reported whether they were the biological mother of the child (yes/no). Additional variables included livelihood (urban/peri-urban, pastoral, agropastoral, fisherfolk), survey zone (East, North, South, West), and survey wave, which served as a proxy for seasonality. Shocks were categorized as climatic, economic, biological, or conflict, and analyzed as the total number across categories (0-1, 2, 3, 4). Child-specific variables, such as age and sex, were considered.

### Data analysis

To guide our analysis, we developed an analytic framework **(Figure 1: Annex A)** that aligns with the conceptual framework for acute malnutrition in Africa’s drylands (H. Young, 2020). For each level, we selected variables that most accurately captured key drivers outlined in the theoretical model. We then added other variables based on their theoretical importance given the current literature, parsimonious representation of key concepts (e.g., use of a validated scale for food insecurity or indicators that capture multiple exposures), and their associations with the outcome variable.

**Figure 1:**
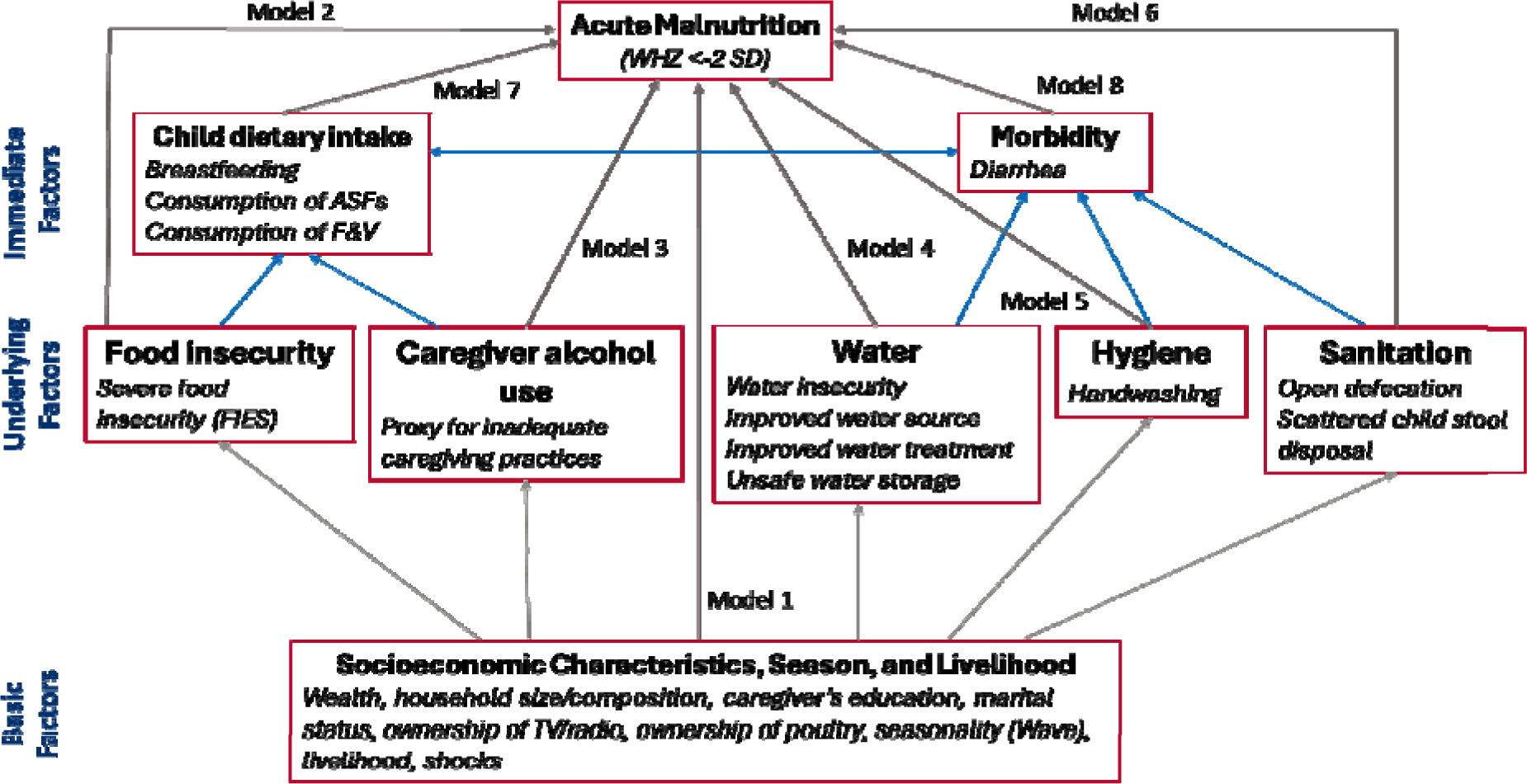
Annex A: Analytical framework of drivers of acute malnutrition illustrating modelling pathways

We used generalized estimating equations to assess the associations of immediate, underlying, and basic factors with acute malnutrition based on the pooled data from all six survey waves. The generalized estimating equations assumed a binomial distribution, a logit link function, an exchangeable correction matrix, and a robust variance estimator. After initial bivariate analyses to identify variables to include in multivariable analyses using a cutoff of P<0.20, we built eight models to identify factors associated with acute malnutrition at P<0.05 (**Figure 1**). In model 1, we assessed the associations between basic risk factors and acute malnutrition. In model 2, we examined the association between food insecurity and acute malnutrition. Model 3 examined the association of caregiver alcohol consumption with acute malnutrition. Models 4 to 6 examined the association between the water, sanitation, and hygiene variables with acute malnutrition. Models 7 and 8 examined the associations of dietary intake variables and diarrhea, respectively, with acute malnutrition. Models 2 to 8 adjusted for the same basic factors, including survey zone, livelihood zone, survey wave, and child’s age and sex. Furthermore, we built a distinct breastfeeding model using data from children under three years old to assess the impact of breastfeeding on this specific age group. Data were analyzed separately for each county using Stata 18 (StataCorp, College Station, Texas, USA).

## Results

### Characteristics of the samples

At Wave 1, the prevalence of acute malnutrition was 21.8% in Turkana and 23.3% in Samburu (**Table 1: Annex E**). In Turkana, 54.8% of the children were male, while in Samburu, 47.8% were male. Across counties, a plurality of children were 11 months old or younger (45.2% in Samburu, 39.7% in Turkana) Most caregivers had no formal education, were 25-34 years old, lived in male-headed households, and resided in pastoral zones.

**Table 1:** Annex E: Child, caregiver, household, and community characteristics at Wave 1 by county

| Characteristic | Turkana (N=1211)<br>% | Samburu (N=586)<br>% |
| --- | --- | --- |
| **Characteristics of children** |  |  |
| Prevalence of acute malnutrition | 21.8 | 23.3 |
| Gender |  |  |
| Male | 54.8 | 47.8 |
| Female | 45.2 | 52.2 |
| Age (Months) |  |  |
| 0-11 | 39.7 | 45.2 |
| 12-23 | 30.8 | 33.3 |
| 35+ | 32.6 | 14.7 |
| **Characteristics of caregivers/mothers** |  |  |
| Age (years) |  |  |
| <25 | 19.8 | 38.5 |
| 25-34 | 47.6 | 46.8 |
| 35 and above | 32.6 | 14.7 |
| Marital status |  |  |
| Married only wife | 50.1 | 54.2 |
| Unmarried | 14.8 | 14.3 |
| Married co-wife | 35.1 | 31.4 |
| Gravidity |  |  |
| <3 | 23.6 | 36.4 |
| 3-5 | 51.2 | 33.2 |
| 5 and above | 25.1 | 30.3 |
| Parity |  |  |
| Less than 2 | 25.9 | 33.7 |
| 3-5 | 49.6 | 34.0 |
| 6 or more | 24.4 | 32.3 |
| Highest level of education |  |  |
| Formal education | 13.6 | 19.6 |
| No formal education | 86.4 | 80.4 |
| **Characteristics of households** |  |  |
| Age of the household head (years) |  |  |
| <25 | 10.8 | 11.3 |
| 25-34 | 34.5 | 36.3 |
| 35 and above | 54.6 | 52.4 |
| Sex of household head |  |  |
| Male | 61.9 | 83.7 |
| Female | 38.1 | 16.3 |
| Household wealth tertile |  |  |
| Lowest | 45.6 | 55.7 |
| Middle | 33.8 | 30.5 |
| Highest | 20.6 | 13.8 |
| Household size |  |  |
| < 4 | 9.6 | 11.7 |
| 4 - 6 | 49.5 | 53.5 |
| 7+ | 40.9 | 34.8 |
| **Community factors** |  |  |
| Livelihood |  |  |
| Urban/peri-urban | 6.5 | 11.4 |
| Pastoral | 63.9 | 83.4 |
| Agropastoral | 14.8 | 5.2 |
| Fisherfolk | 14.8 | na. |
| Survey zone |  |  |
| Central | 21.0 | 7.1 |
| North | 25.6 | 77.1 |
| South | 24.1 | na. |
| East | na. | 15.8 |
| West | 29.2 | na. |

<sup>1</sup>These variables are considered as basic or systemic factors in the analysis

### Trends in acute malnutrition by county and livelihood

Acute malnutrition prevalence among children under 5 was persistently high across survey waves, with no significant changes from Wave 1 to Wave 6 (**Figure 2: Annex B**). When examined by livelihood zone, acute malnutrition decreased from Wave 1 to Wave 6 among pastoralists (20.7% to 18.2%), while there was an increase among agro-pastoralists (19.1% to 27.3%), fisherfolk (26.5% to 31.5%), and urban/peri-urban dwellers (27.6% to 29.5%) in Turkana (**Figure 3: Annex C**). In Samburu, the prevalence of acute malnutrition remained persistently high, showing no improvement in trends between Waves 1 and 6 (**Figure 3)**. Although the prevalence in the agropastoral and urban/peri-urban livelihoods was lower than among pastoralists, there was a general increase in the trends of acute malnutrition between Waves 1 and 6.

**Figure 2:**
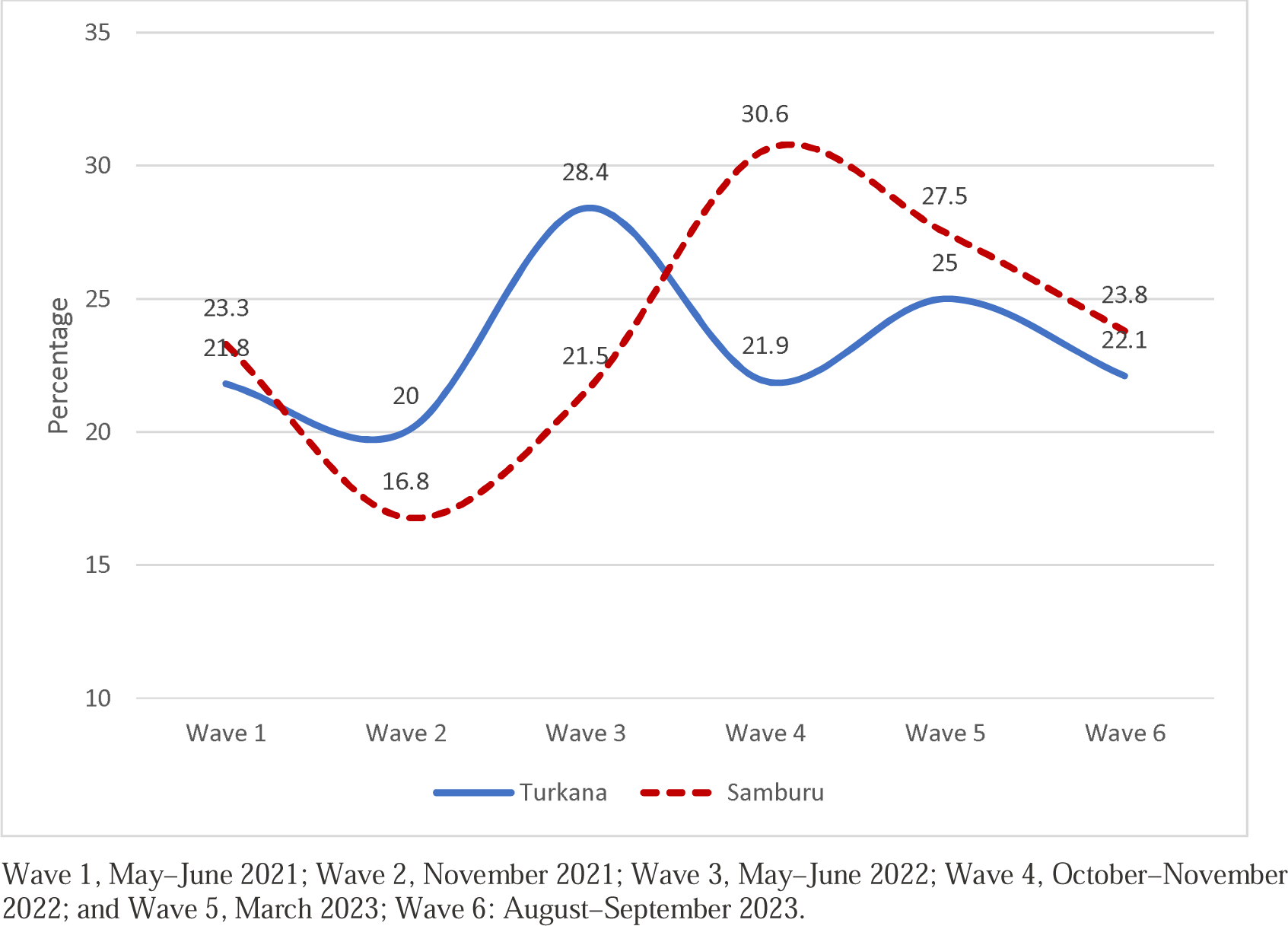
Annex B: Trends in the prevalence of acute malnutrition (WHZ < -2 SD) among children by county, Turkana and Samburu

**Figure 3:**
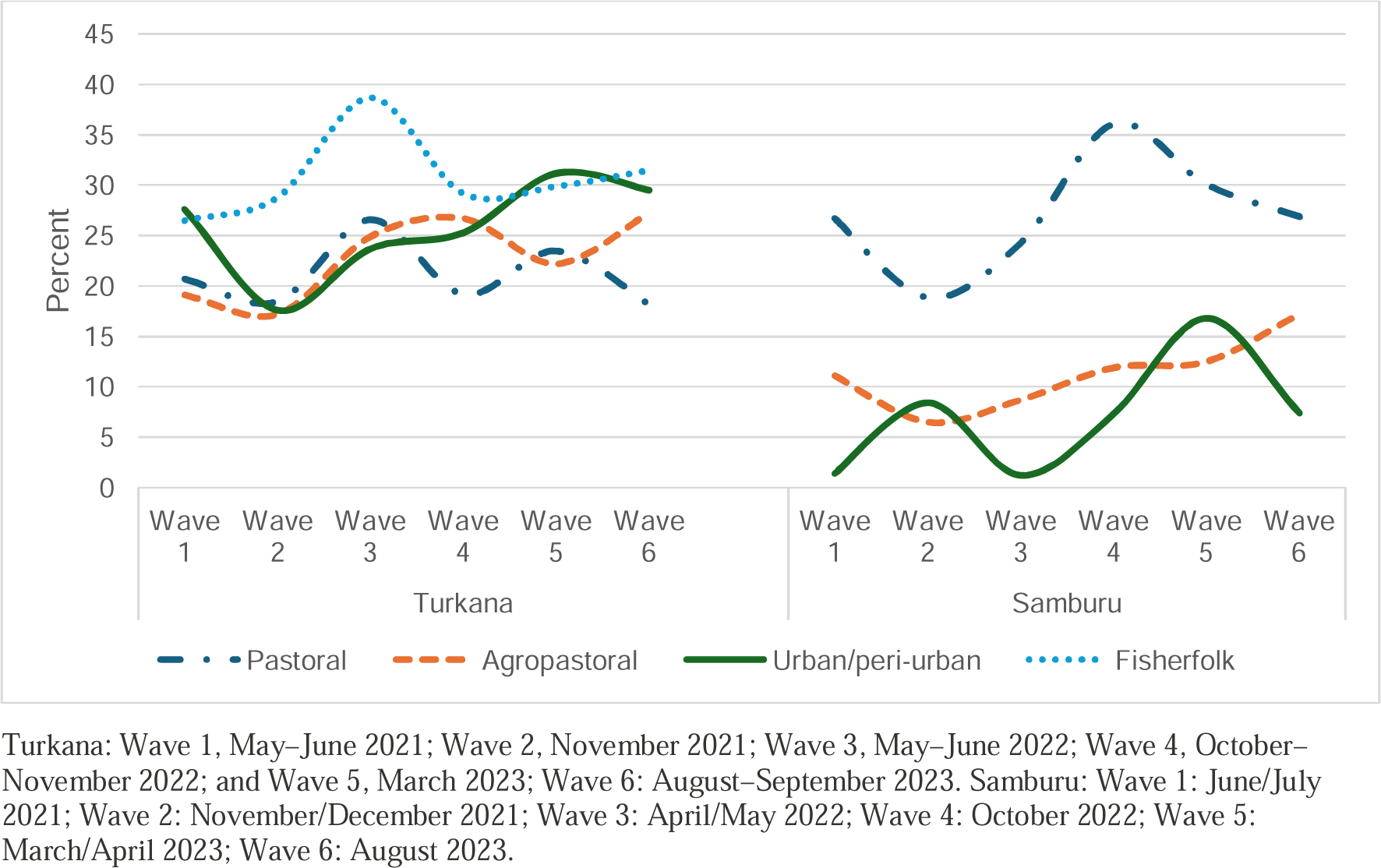
Annex C: Trends in the prevalence of acute malnutrition (WHZ < -2 SD) among children by livelihood and county

### Basic risk factors of acute malnutrition

In multivariable models of basic factors, female children, compared with male children, were 21% (aOR=0.79, 95% CI=0.65, 0.97) lower odds of acute malnutrition in Turkana and 41% (aOR=0.59, 95% CI=0.42, 0.82) lower odds in Samburu (**Table 2: Annex F**). Child age was not associated with acute malnutrition. In Samburu, but not Turkana, children of caregivers with formal education had 44% (aOR=0.56; 95% CI=0.35, 0.88) lower odds of acute malnutrition than those whose caregivers did not have any formal education. In Turkana only, compared with children of caregivers younger than 20 years, children of caregivers aged 20-34 or 35 years or older had 38% (aOR=0.62; 95% CI=0.42, 0.92) and 42% (aOR=0.58; 95% CI=0.38, 0.90) lower odds of acute malnutrition, respectively. Additionally, children of caregivers with 3-5 or 6 or more children had 39% (aOR=1.39; 95%CI=1.12, 1.73) and 70% (aOR=1.70; 95%CI=1.30, 2.23) higher odds of acute malnutrition, respectively, compared with those with 2 or fewer children. Compared with children in male-headed households, children in female-headed households had 34% (aOR=1.34; 95%CI=1.07, 1.66) and 81% (aOR=1.81; 95%CI=1.07, 3.07) higher odds of acute malnutrition in Turkana and Samburu, respectively. Additionally, children in the fisherfolk livelihood in Turkana had 36% (aOR=1.36; 95%CI=1.01, 1.82) higher odds of acute malnutrition compared with those in urban or peri-urban areas. The odds of acute malnutrition were 15% (aOR=0.85; 95%CI=0.72, 1.00) and 29% (aOR=0.71; 95%CI=0.53, 0.94) lower at Wave 2 than at Wave 1 in Turkana and Samburu, respectively.

**Table 2:** Annex F: Associations between basic factors and acute malnutrition among <u>children under five years of age in Turkana and Samburu</u>

| Variable | Turkana (N=1201)<br>aOR [95% CI] | Samburu (N=582)<br>aOR [95% CI] |
| --- | --- | --- |
| **Characteristics of children** |  |  |
| Female child (ref: male) | 0.79 [0.65, 0.97]* | 0.59 [0.42, 0.82]** |
| Child's age (ref: <1 year) |  |  |
| 1 year | 1.11 [0.91, 1.36] | 0.84 [0.61, 1.16] |
| 2 years | 1.01 [0.78, 1.30] | 0.85 [0.57, 1.28] |
| 3 years | 1.06 [0.77, 1.48] | 1.43 [0.84, 2.41] |
| 4 years | 1.38 [0.92, 2.07] | 1.83 [0.96, 3.49] |
| **Characteristics of caregivers/mothers** |  |  |
| Caregiver received formal education (ref: did not) | 0.95 [0.76, 1.19] | 0.56 [0.35, 0.88]* |
| Caregiver age (ref: <20 years) |  |  |
| 20-34 years | 0.62 [0.42, 0.92]* | 0.77 [0.55, 1.08] |
| 35+ years | 0.58 [0.38, 0.90]* | 0.72 [0.47, 1.10] |
| Caregiver marital status (ref: married, only wife) |  |  |
| Unmarried | 0.89 [0.73, 1.08] | 1.11 [0.72, 1.72] |
| Married, co-wife | 0.86 [0.70, 1.07] | 1.07 [0.73, 1.57] |
| Caregiver biological mother (ref: not) | 1.02 [0.77, 1.35] | 1.27 [0.69, 2.34] |
| Caregiver number of children (ref: 0-2 children) |  |  |
| 3-5 | 1.39 [1.12, 1.73]** | 0.94 [0.65, 1.36] |
| 6 or more | 1.70 [1.30, 2.23]*** | 1.15 [0.75, 1.76] |
| **Characteristics households** |  |  |
| Female household head (ref: male) | 1.34 [1.07, 1.66]** | 1.81 [1.07, 3.07]* |
| Household owns poultry (ref: does not) | 1.10 [0.91, 1.33] | 1.00 [0.68, 1.48] |
| Household owns TV or radio (ref: does not) | 0.92 [0.71, 1.19] | 0.89 [0.65, 1.22] |
| Wealth tertile (ref: highest) |  |  |
| Lowest | 1.19 [0.94, 1.51] | 0.98 [0.61, 1.58] |
| Middle | 1.03 [0.84, 1.27] | 1.00 [0.68, 1.48] |
| Shocks (ref: 0-1 shocks) |  |  |
| 2 shocks | 0.98 [0.79, 1.23] | 0.80 [0.59, 1.07] |
| 3 shocks | 0.91 [0.73, 1.14] | 0.75 [0.55, 1.01] |
| 4 shocks | 0.99 [0.78, 1.25] | 0.74 [0.53, 1.05] |
| **Community factors** |  |  |
| Survey zone (ref: central) |  |  |
| East | na. | 0.86 [0.52, 1.40] |
| North | 1.12 [0.80, 1.58] | 1.30 [0.81, 2.09] |
| South | 1.15 [0.88, 1.50] | na. |
| West | 0.88 [0.67, 1.18] | na. |
| Livelihood zone (ref: urban/peri-urban) |  |  |
| Pastoral | 0.88 [0.76,1.03] | 1.30 [0.93,1.82] |
| Agropastoral | 0.83 [0.67,1.03] | 1.11 [0.80,1.54] |
| Fisherfolk | 1.36 [1.01,1.82]* | na. |
| Survey wave (ref: Wave 1) |  |  |
| Wave 2 | 0.85 [0.72, 1.00]* | 0.71 [0.53, 0.94]* |
| Wave 3 | 1.16 [0.96, 1.39] | 0.82 [0.59, 1.12] |
| Wave 4 | 0.89 [0.72, 1.10] | 1.11 [0.79, 1.57] |
| Wave 5 | 1.00 [0.80, 1.27] | 0.96 [0.65, 1.42] |
| Wave 6 | 0.78 [0.59, 1.02] | 0.86 [0.55, 1.33] |

\* p<0.05, \*\* p<0.01, \*\*\* p<0.001; aOR = adjusted odds ratio; 95% CI = 95% confidence interval; na = not applicable

### Immediate and underlying risk factors of acute malnutrition

Children who experienced diarrhea in the two weeks before the surveys, compared with those who did not, had 19% (aOR=1.19, 95%CI=1.06, 1.34) and 23% (aOR=1.23, 95%CI=1.02, 1.47) higher odds of acute malnutrition in Turkana and Samburu, respectively (**Table 3: Annex G**). Children who consumed animal-source foods in the 24 hours before the surveys, compared with those who did not, had 12% (aOR=0.88, 95%CI=0.78, 0.99) and 22% (aOR=0.78, 95%CI=0.65, 0.93) lower odds of acute malnutrition in Turkana and Samburu, respectively. In Samburu, the odds of acute malnutrition were 38% (aOR=1.38, 95%CI=1.05, 1.82) higher in children who received breastmilk compared with those who did not; this association was not significant when restricted to children younger than 3 years (*results not shown)*. In Turkana only, children of caregivers who used alcohol had 32% (aOR=1.32, 95%CI=1.10, 1.60) higher odds of acute malnutrition compared with those whose caregivers who did not. Household food insecurity and the water, sanitation, and hygiene variables (household water insecurity, caregiver hand washing, and open defecation) were not associated with acute malnutrition in either county.

**Table 3:** Annex G: Associations of immediate and underlying factors with acute malnutrition among children under five years of age in Turkana and Samburu

|  | **Turkana (N=1201)** | **Samburu (N=582)** |
| --- | --- | --- |
| **Variables** | **aOR [95% CI]** | **aOR [95% CI]** |
| *Immediate factors* |  |  |
| Child diarrhea (ref: no diarrhea) | 1.19 [1.06, 1.34]** | 1.23 [1.02, 1.47]* |
| Consumed animal-source foods (ref: did not) | 0.88 [0.78, 0.99]* | 0.78 [0.65, 0.93]** |
| Consumed fruits and vegetables (ref: did not) | 0.88 [0.76, 1.01] | 1.14 [0.93, 1.40] |
| Consumed breastmilk (ref: did not) | 1.11 [0.96, 1.29] | 1.38 [1.05, 1.82]* |
| *Underlying Factors* |  |  |
| Moderate-to-severe food insecurity (ref: no-to-mild food insecurity) | 1.00 [0.83, 1.21] | 1.01 [0.75, 1.34] |
| Caregiver consumes alcohol (ref: does not) | 1.32 [1.10, 1.60]** | 1.33 [0.81, 2.20] |
| Household water insecurity (ref: no-to-marginal) |  |  |
| Low | 1.05 [0.89, 1.23] | 0.97 [0.74, 1.26] |
| Moderate | 1.03 [0.90, 1.19] | 0.89 [0.69, 1.16] |
| High | 1.04 [0.88, 1.23] | 1.12 [0.80, 1.57] |
| Caregiver wash hands after using toilet (ref: does not wash hands) | 1.08 [0.94, 1.24] | 1.11 [0.92, 1.33] |
| Open defecation (ref: no open defecation) | 1.13 [0.96, 1.33] | 1.16 [0.89, 1.53] |

\* p<0.05, \*\* p<0.01, \*\*\* p<0.001; aOR = adjusted odds ratio; 95% CI = 95% confidence interval

## Discussion

This longitudinal study examined factors associated with acute malnutrition among children under five in two arid and semi-arid counties in Kenya. Our findings reveal persistently high levels of acute malnutrition over the 24-month study period, with no improvement and even worsening trends among households in several livelihoods. The descriptive analysis revealed that acute malnutrition remained high in Turkana and Samburu, showing no improvement over the 24-month study period. These results, and prior work in this region (Ministry of Health, 2023), suggest that acute malnutrition remains a priority issue in this area despite concerted efforts by government and development partners to intervene. Additionally, we identified factors associated with both increased and reduced risk of acute malnutrition, findings that have practical implications for nutrition and multi-sectoral interventions in these counties.

Among the examined basic factors, we found that children residing in households headed by women were at a higher risk of experiencing acute malnutrition compared with those living in households headed by men. While women often prioritize food security and dietary diversity, positively impacting children’s nutrition (Addai, Ng’ombe, & Temoso, 2022; Smith, Ramakrishnan, Ndiaye, Haddad, & Martorell, 2003), financial constraints and limited access to resources in this setting may hinder their ability to provide adequate nutrition, as previously found in Malawi (Chirwa & Ngalawa, 2008). Children of caregivers with more than three children also had higher odds of acute malnutrition. Larger families may face challenges as household members must share limited resources, potentially resulting in smaller meals and inadequate nutrition (Shafiq et al., 2021). These findings underscore the need for targeted interventions to support female-headed households and larger families, which could significantly improve children’s nutritional status in these vulnerable groups.

Caregiver education was another important basic factor associated with acute malnutrition. In Samburu, children of caregivers with formal education had a lower risk of acute malnutrition compared with those whose caregivers who lacked formal education, which aligns with prior literature (Abdirahman, Chege, & Kobia, 2019; Ejike, 2016; Iftikhar, Bari, Bano, & Masood, 2017). Formal education provides caregivers with knowledge and skills for informed decisions on child-feeding practices, hygiene, and healthcare utilization (Fadare, Amare, Mavrotas, Akerele, & Ogunniyi, 2019). The influence of caregiver education on child nutrition may, however, vary based on the local context and availability of other sources of nutrition-related knowledge. Indeed, prior research has found that maternal education and nutrition knowledge are distinct predictors of child nutritional status (Webb & Block, 2004), suggesting that targeted nutrition-education interventions in areas with limited formal education, such as Turkana and Samburu, may help to reduce the burden of acute malnutrition (Fadare et al., 2019).

Livelihood was another salient predictor of child acute malnutrition. In Turkana, children in the fisherfolk livelihood had higher odds of acute malnutrition compared with those living in urban or peri-urban areas. Although fish are a nutrient-dense food often associated with improved child nutrition, fishing households may not have equitable access to benefits of their catch, potentially due to a lack of investment and technical support (Fiorella et al., 2014). Further, the reliance on fishing as a livelihood strategy may divert time and resources away from other food-production activities that support child nutritional well-being (Nyawade, Were-Kogogo, Owiti, Osimbo, & Daniel, 2021). To overcome these challenges and fully harness the benefits of fishing as a livelihood strategy, it is essential to implement carefully designed policies and programs that address the unique challenges faced by fishing communities in Turkana. This could involve targeted investments in infrastructure, such as improving storage facilities and access to clean water and enhancing market access for fisherfolk. Capacity-strengthening programs to equip fishing communities with the necessary knowledge and skills for sustainable fishing practices and diversified livelihood opportunities could also be beneficial. Furthermore, addressing the underlying social, economic, and environmental factors that affect food security and nutritional outcomes in the region is crucial. This may entail initiatives to strengthen social safety nets and efforts that support food production to improve the nutritional outcomes of children in the fisherfolk zone.

Among the examined immediate factors, diarrhea was associated with higher odds of acute malnutrition. The well-established bidirectional relationship between malnutrition and diarrhea indicates that interventions must address both aspects simultaneously (Brown, 2003). Diarrheal diseases lead to fluid and electrolyte losses, compromised nutrient absorption, reduced appetite, and increased nutrient demands, all contributing to malnutrition (Brown, 2003; Hodges et al., 2015; Weisz et al., 2011). In the other direction, malnutrition impairs immune function, increasing susceptibility to diarrhea (Brown, 2003). Addressing the impact of diarrhea on acute malnutrition requires a multifaceted approach, combining improvements in water, sanitation, and hygiene practices, appropriate treatment and management of diarrhea, and interventions to enhance dietary intake and overall nutritional status (Mwaniki & Makokha, 2013; Nounkeu et al., 2021).

Certain aspects of a child’s diet were also significantly associated with acute malnutrition. In both counties, animal-source food consumption was associated with lower odds of acute malnutrition among children under five. Numerous studies have demonstrated that animal-source foods provide high-quality proteins, vitamins, and minerals essential for child growth and development (Chege, Kimiywe, & Ndungu, 2015; Dror & Allen, 2011; White et al., 2021). As such, interventions that promote animal-source food consumption can potentially prevent acute malnutrition and promote children’s overall health (Balehegn, Mekuriaw, Miller, McKune, & Adesogan, 2019; White et al., 2021).

Concerning underlying factors, we found that caregiver alcohol consumption was associated with higher odds of child acute malnutrition. Alcohol use can restrict caregivers’ abilities to provide adequate care, resulting in impaired judgment, reduced responsiveness, and decreased ability to create a supportive environment that young children need for healthy growth and development (Phillips & Adams, 2001). Children of caregivers who use alcohol may also be at a higher risk of experiencing neglect, abuse, or instability at home, which can further compromise their well-being (Burchinal, 1999). Efforts to reduce caregiver alcohol use for example through screening and community-based alcohol brief interventions implemented by community health workers (Takahashi et al., 2018), may reduce the burden of acute malnutrition in Turkana.

Unexpectedly, food insecurity and water, sanitation, and hygiene variables were not associated with acute malnutrition in either county. It is possible that the basic factors that influence food insecurity also impact diet, masking a potential direct association when all factors are examined concurrently. Limited variability in water, sanitation, and hygiene practices may have precluded the detection of associations, despite water, sanitation, and hygiene being critical issues in these counties.

### Study strengths and limitations

A major strength of this study was its longitudinal design, which allowed us to examine which risk factors of malnutrition are most important during critical periods of child development. Additionally, we used generalized estimating equations, which allowed us to account for intra-individual correlation, assessed numerous factors across different levels of the framework, and had large sample sizes with little attrition in both counties.

However, the study has limitations. Although the attrition rate in the study was low, it could compromise the robustness of some estimates. Furthermore, considering the nature of the analysis, we could not establish causality between the study variables. Thus, our conclusions are carefully restricted to statements about the associations between the explanatory and outcome variables. This study was designed to capture seasons by collecting data during different expected seasons. However, Waves 2-6 were considered a drought period with successive failed rains; therefore, the seasons were not as expected, thus limiting our ability to connect different factors that could contribute to acute malnutrition with season. Despite these limitations, understanding the factors associated with acute malnutrition in Turkana and Samburu regions over time is crucial for effective intervention and program design.

## Conclusions

Our study highlights persistently high levels of acute malnutrition in Turkana and Samburu, Kenya, with several factors associated with greater risk, including female-headed households, larger family size, lack of caregiver education, livelihood zone, caregivers’ alcohol use, and recent experience of diarrhea. Conversely, child consumption of animal-source foods was associated with lower odds of acute malnutrition. Gender-specific interventions and programs could be implemented to address acute malnutrition in these vulnerable households. Targeted interventions to reduce child diarrhea and caregiver alcohol use and to increase consumption of animal-source foods could also help reduce acute malnutrition. Our findings underscore the need for multifaceted interventions that concurrently address multiple underlying, basic, and immediate factors to effectively reduce acute malnutrition in these and similar settings.

## Ethics statement

The African Population and Health Research Center (APHRC) obtained ethical and research approvals and research permits from Amref Health Africa’s Ethical and Scientific Review Committee (Amref ESRC P905/2020) and the National Commission for Science, Technology, and Innovation of Kenya and signed a reliance agreement with RTI International’s Institutional Review Board for the research.

## Data sharing

Data from this study are available to researchers through the APHRC’s Microdata Portal (https://aphrc.org/microdata-portal/).

## Contributor Statement

DAA, ES, FT, VLF, CW, BM, EA, HO, SE, GC, WA, NM, EK, CL and BS performed the research. DAA, ES, VLF, CL, FT and BS designed the research study. CW, WA, BM, EA, HO, SE, EK, GC and NM contributed essential reagents or tools. LA, JDM and BM analyzed the data. DAA wrote the first draft of the paper. DAA, ES, FT, VLF, CW, EK, CL, LA and JDM critically reviewed the manuscript. All authors have read and approved the final manuscript.

## Data Availability

Data from this study are available to researchers through the APHRC Microdata Portal.

https://aphrc.org/microdata-portal/

---

**Supplemental Figure 1:**
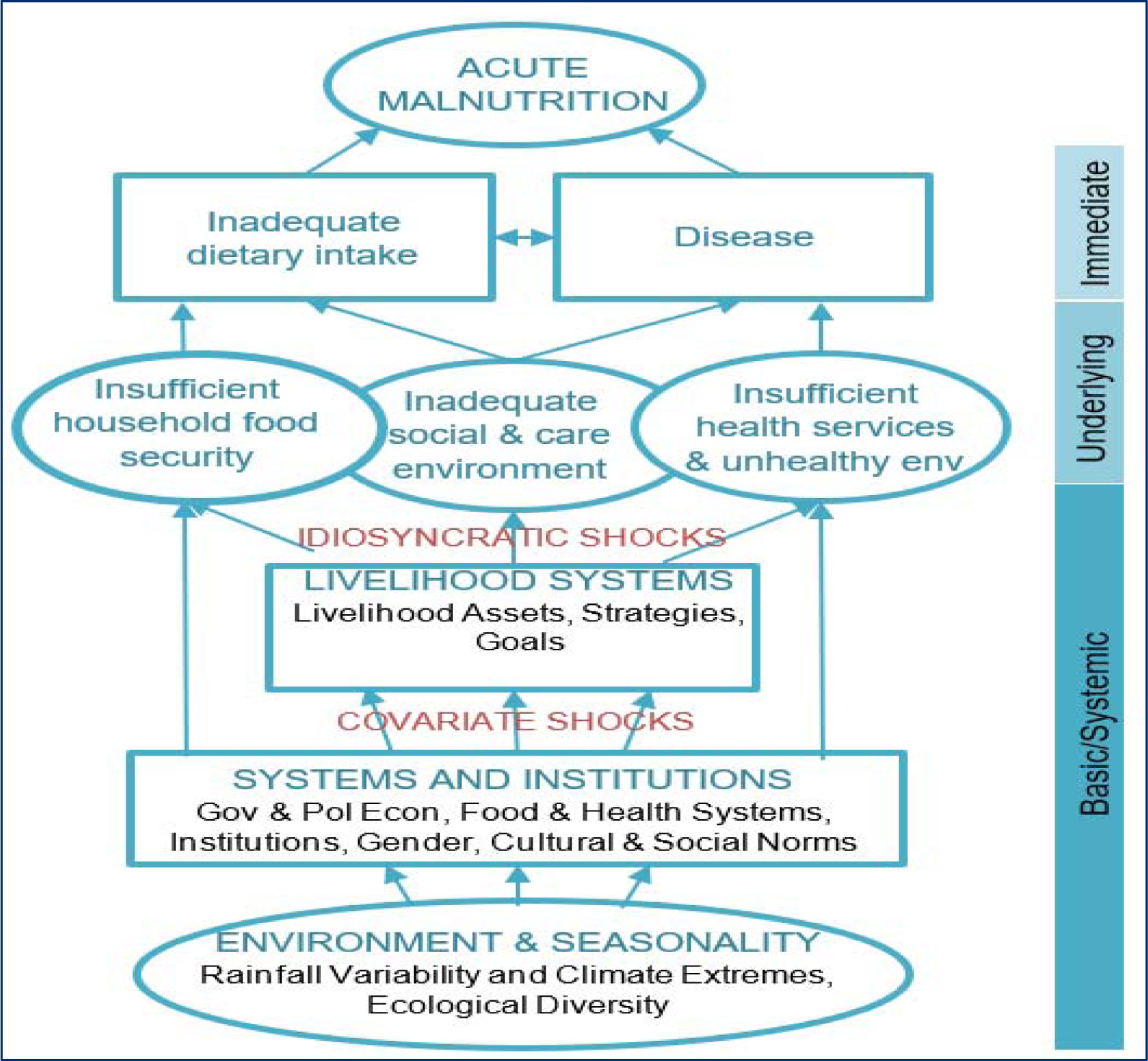
Annex D: A conceptual framework for acute malnutrition in Africa’s drylands (Young, 2020)

